# Establishing a baseline for human cortical folding morphological variables: a multicenter study

**DOI:** 10.1101/2022.03.10.22272228

**Authors:** Fernanda Hansen Pacheco de Moraes, Victor B. B. Mello, Fernanda Tovar-Moll, Bruno Mota

## Abstract

Differences in the way human cerebral cortices fold have been correlated to health, disease, development, and aging. But to obtain a deeper understating of the mechanisms that generate such differences it is useful to derive one’s morphometric variables from first principles. This work explores one such set of variables that arise naturally from a model for universal self-similar cortical folding that was validated on comparative neuroanatomical data. We aim to establish a baseline for these variables across the human lifespan using a heterogeneous compilation of cross-sectional datasets, as the first step to extend the model to incorporate the time evolution of brain morphology. We extracted the morphological features from structural MRI of 3650 subjects: 3095 healthy controls (CTL) and 555 Alzheimer’s Disease (AD) patients from 9 datasets, which were harmonized with a straightforward procedure to reduce the uncertainty due to heterogeneous acquisition and processing. The unprecedented possibility of analyzing such a large number of subjects in this framework allowed us to compare CTL and AD subjects’ lifespan trajectories, testing if AD is a form of accelerated aging at the brain structural level. After validating this baseline from development to aging, we estimate the variables’ uncertainties and show that Alzheimer’s Disease is similar to premature aging when measuring global and local degeneration. This new methodology may allow future studies to explore the structural transition between healthy and pathological aging and may be essential to generate data for the cortical folding process simulations.

**Significance Statement:** Understating Cortical folding is of increasing interest in neurosciences as it has been used to discriminate disease in humans while integrating pieces of knowledge from compared neuroanatomy and neuroproliferations programs. Here we propose estimating the baseline of cortical folding variables from multi-site MRI human images, evaluating the changing rate of its independent variables through the human lifespan, and proposing a simple harmonization procedure to combine multicentric datasets. Finally, we present a practical application of these techniques comparing Alzheimer’s Disease and Cognitive Unimpaired Controls based on the estimated changing rates.

**Highlights:**

- Baseline of independent cortical folding variables from 3650 multi-site human MRI
- Propose a simple harmonization procedure to combine multicentric datasets
- Evaluate the changing rate of independent variables through the human lifespan
- Practical application comparing Alzheimer’s Disease and Controls rates

## Introduction

Mapping the human brain development and aging longitudinally from a unique dataset is barely impossible due to our extensive lifespan. Suppose one wants to understand the time evolution of the brain morphology through the whole lifespan. In this case, it is mandatory to combine multiple datasets and devise methods that allow a fair comparison among them. The recent advent of large heterogeneous datasets (to ensure the legit results across several populations) and data curation innovations allowed neuroscience to explore the brain’s changes on an enormous scale, from functional data to structural studies. Here, we propose a novel methodology of combining structural MRI from different acquisition sites and equipment, acknowledging their heterogeneity and providing insights about cortical folding during development and aging, with a practical application of these results in Alzheimer’s Disease, the most common dementia worldwide. This effort is essential to provide experimental information about the evolution span of the cortex morphology and build a cortical folding theory.

In the past years, the study of cortical folding in humans and other mammals was extensively promoted by the application of translational science, open-access databases, and data science tools which allowed the field to grow in multiple and sometimes convergent directions [1–4]. Due to its biological background, cortical folding measurements have been included in human brain structure analysis, discriminating disease from healthy controls [5, 6], describing its correlation with cognition [7], and relation to aging [8, 9]. Despite the lack of consensus defining a unique theory that explains cortical folding in every scale, Mota & Herculano-Houzel [10] proposed a cortical folding model respected by more than 50 mammals brain hemispheres. It predicts a power-law relationship between cortical thickness (T), exposed (A_*E*_), and total area (A_*T*_) (Equation 1).

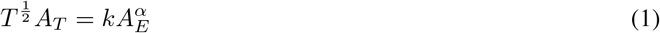

Where *α* is the fractal index and universal constant, with a theoretical value of 1.25 and calculated as 1.305 [10] when considering a heterogeneous dataset of different species, thus, the usual geometrical variables used to describe the cortex are not independent in this model.

Wang et al. continued this work, validating the theory for different groups of humans [11] in specific regions of interest [12], obtaining new evidence that the cortex is indeed a fractal structure. In 2021, Wang et al. showed that from this model one could derive a more natural set of nearly independent morphological variables, K, S, and I, that could be used to improve disease discrimination from regular and typical structural changes of the brain, like aging [13].

In this framework, the morphology of each cortex is expressed as a point in a three-dimensional abstract space, with each coordinate component corresponding to the log-value of an independent morphometric variable. The use of log-values guarantees that linear combinations of the basis vectors correspond to power-law relations. Conversely, as long as any new variables are expressible as power laws, different but equivalent sets of variables can thus be related to each other by a change of Cartesian coordinate bases.

As a starting point, we use the log-values of the commonly morphometric variables, the total area log_10_ A_T_, exposed area log10 A_E_ and average thickness log10 T^2^. We then derive our new variables thusly: *K* = *log*_10_k (Equation 2) is a near-invariant quantity obtained by isolating *k* in Equation 1, and is associated with conserved viscoelastic [REF] properties of cortical matter. S (Equation 3), also dimensionless, encapsulates the aspect of brain shape that changes more significantly across cortices, and I (Equation 4), representing brain isometric volume and carries the information about overall cortical size.^1^. Those three variables combined may help distinguish pathological events similar to age effects, as AD.

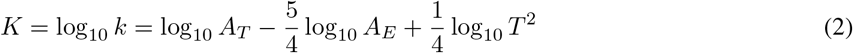

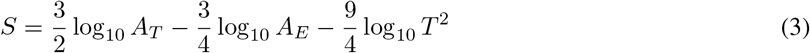

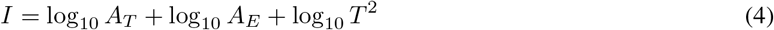

Baseline values of those cortical morphological variables and their inherent uncertainties over the human lifespan were never defined due to methodological limitations of combining multicenter studies of brain structural images [14, 15]. However, both are crucial for applied studies. Uncertainties determine measuring limitations and allow a proper comparison between different cohorts. Baseline values allow clinical applications and, when necessary, simulations of healthy control data. Multi-center studies have become more common in the last few decades due to improved data sharing and the open science trend. However, combining MRI data from multiple experiments is not a trivial task. Part of these uncertainties is explained by type A errors, i.e., random, which appear as a consequence of the natural fluctuation within the species and from the data acquisition itself. Theoretically, these random errors are the same if one considers multiple experiments.

Confounding components could be added when gathering MRI images with differences in acquisition parameters, acquisition equipment, and versions of the post-processing software [15, 16]. Together, these effects can add type B errors to the data, ultimately reflecting a systematic difference in morphological variables calculated from different databases and increasing the data spread. In practice, the natural fluctuations are convoluted with the type B errors, enhancing the data spread.

Recently, a vast literature in primary morphological variables from MR images, [17–19], and more complex ones [14], explored the limitations of compiling images acquired with different scanners and protocols and its implication in statistical analysis results. The suggested solution for these limitations is a procedure called data harmonization, in which there is an “explicit removal of site-related effects in multi-site data” [18].

It is essential to add that one cannot, in general, untangle the contributions of natural fluctuation and data acquisition to random errors. Also, it is not possible to distinguish between type B errors of the acquisition and processing. For this, one would need a diligent procedure to track all differences between acquisition and processing as well as multiple images of the same individual for each sample. This manuscript deals only with the first case, estimating the type A errors due to acquisition and processing using three subsequent structural images of the subjects^2^. The uncertainties associated with the variables (K, S, and I) are summarized in Equation 5.

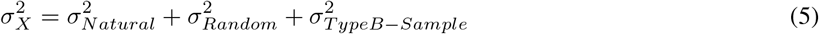

In this manuscript, we developed a simplistic harmonization method that allows a multi-site analysis, determining K, S, and I baseline values, their uncertainties, and the ratio of changing rates through years for cross-sectional images of healthy controls. Although this work focused is these novel morphological variables, we also provided the baseline of usual morphological variables such as thickness *T* and Gyrification Index GI. A robust estimation framework of a baseline time function is important to provide future clinical studies enough information to compare brain structure trajectories in case of a pathological investigation. We then, as a clinical application and extension of de Moraes et al. [20], and Wang et al. [12], verify if Alzheimer’s Disease (AD) is similar to a premature accelerated aging brain in terms of the independent morphological variables, K, S, and I, by comparing their rates. Finally, we have determined the time evolution of the fractal dimension *α* proposed in Mota & Herculano-Houzel’s model [10], interpreting all results within the framework of this theory.

## Materials and Methods

Participants and data included in this analysis were either acquired by the D’Or Institute for Research and Education (IDOR), Rio de Janeiro, RJ, Brazil, used by [20] (approved by the Hospital Copa D’Or Research Ethics Committee under protocol number CAAE 47163715.0.0000.5249), published by Wang et al. [11, 12] or from open-access databases, as AHEAD [21] and AOMIC (PIOP01 and PIOP02) [22]. The total number of subjects is 3095 healthy controls from 4 to 96 years old and 555 Alzheimer’s Disease subjects from 56 to 92 years old. To investigate the type B errors during image acquisition and variances in repeated measures performed in the same conditions, we included the first 50 subjects from the AOMIC ID1000 [22], described in Supplementary Material. AHEAD and HCPr900 only include age range instead of the actual age due to local Ethics Committee rules. To overcome the lack of a exact number, we assumed the interval’s mean age as a reference. Datasets’ demographics are summarized in Table 1.

**Table 1:** Summary for each dataset. Mean value ± standard deviation. Diagnostic: CTL – Healthy control; AD – Alzheimer’s Disease. Details in Supplementary Material, Table 4.

| Datasets | Diagnostic | N (F) | Age (Age range) [years] |
| --- | --- | --- | --- |
| ADNI [23] | CTL | 868 (445) | $75 \pm 6.5$ (56; 96) |
| | AD | 542 (241) | $75 \pm 8$ (56; 92) |
| AHEAD<sup>1</sup> [21] | CTL | 100 (56) | $42 \pm 19$ (24; 76) |
| AOMIC PIOP01 [22] | CTL | 208 (120) | $22 \pm 1.8$ (18; 26) |
| AOMIC PIOP02 [22] | CTL | 224 (128) | $22 \pm 1.8$ (18; 26) |
| HCP900r [24] | CTL | 881 (494) | $29 \pm 3.6$ (22; +36) |
| IDOR | CTL | 77 (53) | $66 \pm 8.4$ (43; 80) |
| | AD | 13 (8) | $77 \pm 6.1$ (63; 86) |
| IXI-Guy’s | CTL | 314 (175) | $51 \pm 16$ (20; 86) |
| IXI-HH | CTL | 181 (94) | $47 \pm 17$ (20; 82) |
| IXI-IOP | CTL | 68 (44) | $42 \pm 17$ (20; 86) |
| NKI [25] | CTL | 168 (68) | $34 \pm 19$ (4; 85) |
| OASIS<sup>6</sup> [26] | CTL | 312 (196) | $45 \pm 24$ (18; 94) |
| **TOTAL** | **CTL** | **3095 (1762)** | **$46 \pm 23$ (4; 96)** |
|  | **AD** | **555 (249)** | **$75 \pm 8$ (56; 92)** |

The structural images from IDOR, AHEAD, AOMIC ID1000, PIOP1, and PIOP2 were processed in FreeSurfer v6.0.0 [27] with the longitudinal pipeline [28] without manual intervention at the surfaces [29]. The FreeSurfer localGI pipeline generates the external surface and calculates each vertex’s local Gyrification Index (localGI) [30]. Values of Average Thickness, Total Area, Exposed Area, and Local Gyrification Index were extracted with Cortical Folding Analysis Tool [31]. We defined as ROI the whole hemisphere, frontal, temporal, occipital, and lateral lobes (based on FreeSurfer definition of lobes). The lobes’ area measurements were corrected by their integrated Gaussian Curvature, removing the partition size effect and directly comparing lobes and hemisphere cortical folding.

Datasets’ demographics, acquisition, and processing information are summarized in Supplementary Material, Table 4.

### Statistical data analysis

The data harmonization procedure is based on the time evolution of the basic morphological variables *T*, A_*T*_ or A_*E*_, modeled as an exponential decay. Using Linear Mixed Models (LMM), we subtracted the sample shift, estimated as the Standard Deviation of Sample Random Effects (log_10_(*Variable*) ∼ *Age* × *ROI* + (1|*Sample* : *ROI*)) in Healthy Controls. From this procedure we directly calculate the variables K, S and I and determine for all morphological variables of interest (also *T*, A_*T*_, A_*E*_,GI = *A*_*t*_*/A*_*e*_) their baseline and uncertainties. This harmonization procedure was validated as the most probable description of the data using the Bayesian comparison framework described in the appendix B. More than that, the Bayesian approach cross-validated all results presented here.

Multiple comparisons of means were made with ANOVA, correlations between cortical folding independent variables, and age estimated with Pearson r and post hoc evaluations with Tukey multiple comparisons of means. The statistical significance threshold was set at *α* = 0.05, and multiple corrections (Bonferroni) were applied when needed. Healthy Control subjects are used as a reference in the necessary normalization and harmonization procedures. A standard linear regression was used to obtain the fractal index *α*.

All analyses were done considering the whole cortical hemisphere and the lobes (see appendix C) as most diseases imply local or nonuniform global structural damage. LMM, standard linear regression and statistics derived from their results were analyzed with RStudio (v1.4.1717 and R v4.1.1). The Bayesian model comparison was developed in Python3 with the package PyMC3.

### Data and code availability

Data from previous publications are available from their repositories: Wang et al., 2016 [11] and Wang et al., 2019 [12]. Data from AHEAD [21] and AOMIC [22] datasets with the cortical folding variables extracted by these authors are available [32]. Data acquired at IDOR does not have clearance for public sharing patients’ information that could lead to identification due to local Ethics Committee approval restrictions but will be shared upon request.

Informed consent was obtained from all subjects for the published datasets as part of the original data acquisition.

## Results

### Multisite harmonization

The analysis starts with data harmonization, a crucial step to combine information from multiple sites allowing a fair comparison among datasets. Figure 1 shows the result of the harmonization procedure and its reflection on the primary morphological variables. The LMMs are used to fit the data assuming a single angular coefficient for all samples and different linear coefficients due to type B errors. We considered the variables trend with age linear based on the correlation between *T*, A_*T*_, and A_*E*_ with Age for the Healthy Control group calculated from our data. There was a significant correlation for T (Pearson r = −0.49; p < 0.0001), A_*T*_ (Pearson r = −0.11; p < 0.0001), and A_*E*_ (Pearson r = −0.05; p < 0.0001). As shown in appendix B, the harmonization was validated as the most probable description of this data.

**Figure 1:**
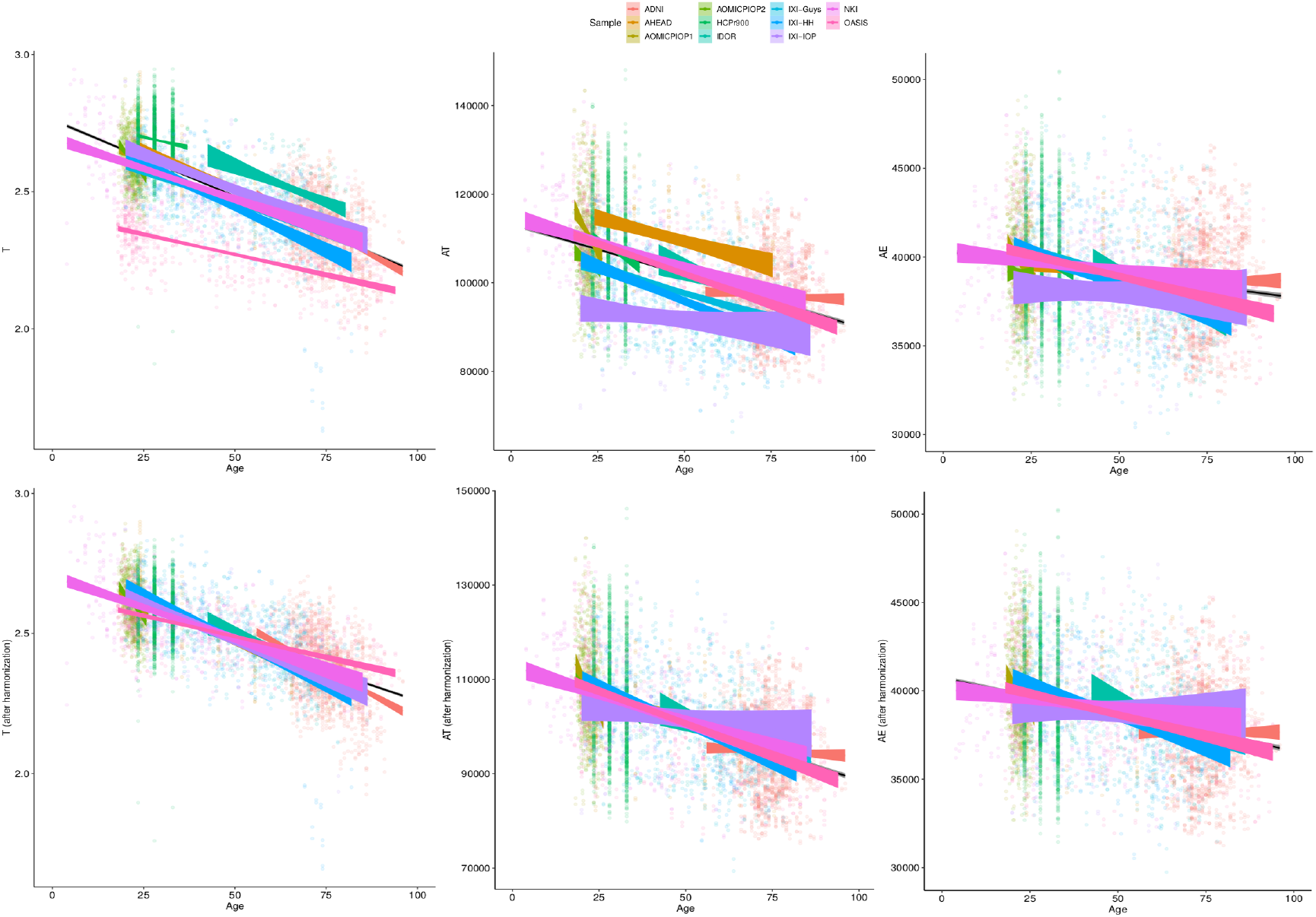
Basic morphological variables through age for Healthy Controls (A) raw data, (B) after harmonizing, removing the estimated residual from the Linear Mixed Model. This procedure results in the harmonized values of the novel morphological variables K, S and I.

**Figure 2:**
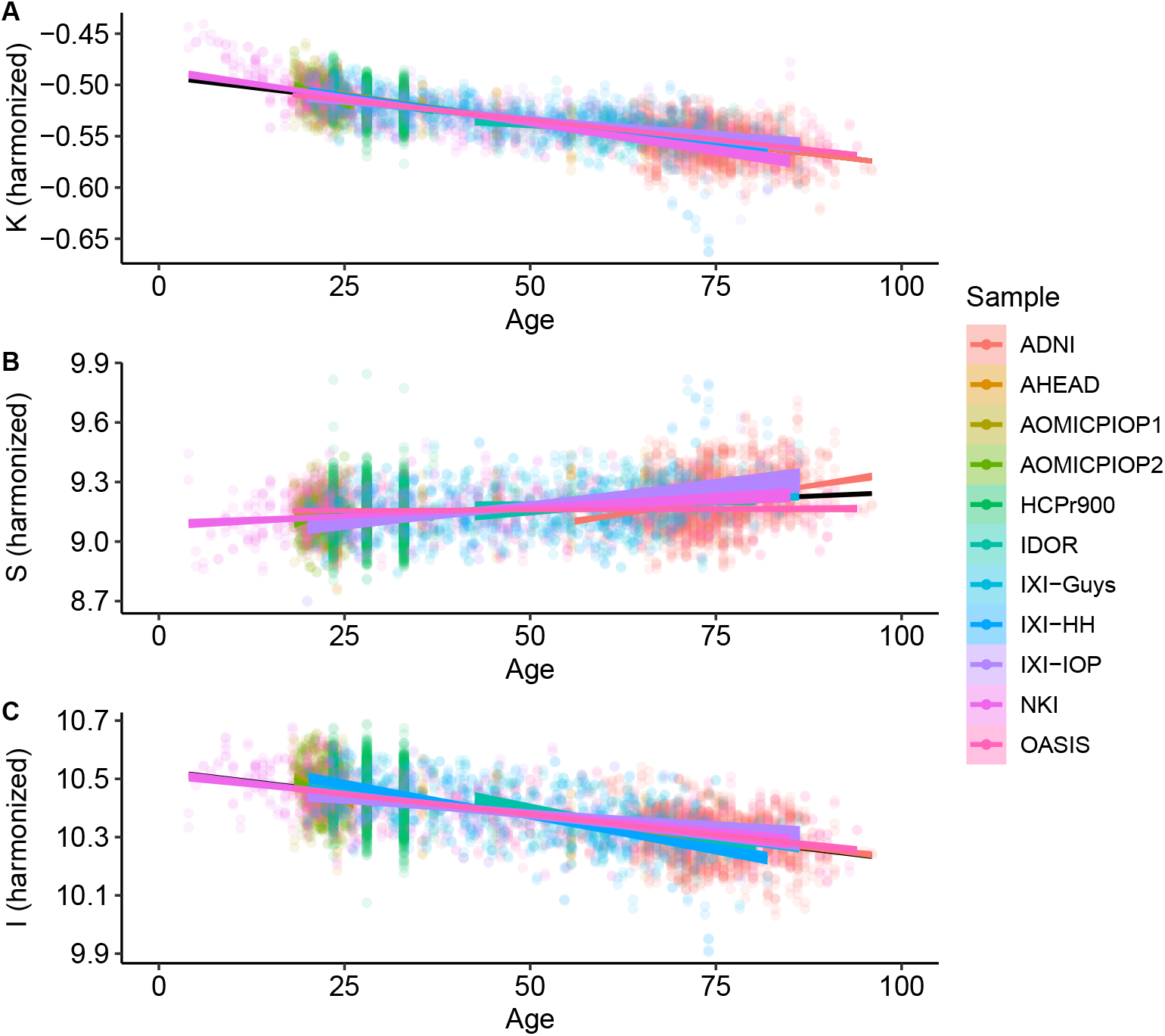
K, S and I through age for Healthy Controls after harmonizing in the primary morphological variables T, A_*T*_, and A_*E*_ (removing the estimated residual from the Linear Mixed Model).

### Baseline, rate estimates, and Alzheimer’s Disease diagnostic discrimination

An immediate consequence of the harmonization is defining a baseline value and its uncertainty for any morphological variable considered in this work. The LMM provide quantitative information about the trends of these variables through age. The estimated uncertainties values are summarized in Table 2 and summaries of linear mixed models are available in Supplementary Material A, Table 5. The so-called Natural Fluctuation represents human diversity and was considered the Residual Standard Deviation of the models. Moreover, we estimated the random component of the experimental error derived from the multiple-image acquisition and processing, described in Supplementary Material C.1. We considered the Random Effect of the Sample category (intercept) Standard Deviation as the type B component of the experimental error. The *σ*_*X*_ is the resultant uncertainty, composed of the previously described errors, and is the square root sum of each one’s squared values. Thereby, *σ*_*K*_ = 0.026 (4.9%), *σ*_*S*_ = 0.14 (1.6%), and *σ*_*I*_ = 0.091 (0.9%) with the corresponding fraction of their healthy controls intercept.

**Table 2:** Summary of the estimated uncertainty components for each morphological variable of interest.

| Variable | Natural Fluctuation | $\sigma_{acquisition} + \sigma_{processing}$ | | $\sigma_X = \sqrt{\sum \sigma_i^2}$ |
| --- | --- | --- | --- | --- |
| | $\sigma_{Natural}$ | $\sigma_{Random}^1$ | $\sigma_{TypeB-Sample}$ | |
| T [mm] | 0.1 | 0.019 | 0.085 | 0.13 (5.35%) |
| $A_T$ [mm<sup>2</sup>] | 9700 | 290 | 4900 | 10871.3 (10.78%) |
| $A_E$ [mm<sup>2</sup>] | 2900 | 70 | 530 | 2948.86 (7.62%) |
| GI | 0.095 | 0.0069 | 0.11 | 0.14 (5.60%) |
| K | 0.016 | 0.0017 | 0.021 | 0.026 (4.94%) |
| S | 0.12 | 0.014 | 0.076 | 0.14 (1.56%) |
| I [mm<sup>3</sup>] | 0.082 | 0.0064 | 0.037 | 0.091 (0.87%) |

<sup>1</sup>repeat measure, mean standard deviation.

The baseline determined by our method is a standard that can be used to compare Health Controls with any atypical group (in terms of brain morphology). It is directly calculated using LMM from the time evolution of the independent morphological variables as shown in 2.

Considering Alzheimer’s Disease, we used the baseline to verify if the age trends of K, S, and I are different between the diseased and Healthy Control groups. We extracted the rates of change for each variable and diagnostic, summarized in Table 3 and displayed in Figure 3. The slopes were compared with post hoc pairwise mean comparisons.

**Table 3:** Changing rate per year for each variable (after harmonization) and diagnostic. Mean value ± standard deviation. Rate [%] was estimated as the fraction from the mean value.

|  | CTL | AD |
| --- | --- | --- |
| T [mm] | $(-440 \pm 6) \times 10^{-5}$ mm/year (-0.18%) | $(-14 \pm 4) \times 10^{-4}$ mm/year(-0.062%) |
| $A_T$ [mm<sup>2</sup>] | $(-240 \pm 5)$ mm<sup>2</sup>/year (-0.24%) | $(-9. \pm 4) \times 10$ mm/year (-0.096%) |
| $A_E$ [mm<sup>2</sup>] | $(-41 \pm 1)$ mm<sup>2</sup>/year (-0.11%) | $(-4 \pm 1) \times 10$ mm/year (-0.1%) |
| GI | $(-350 \pm 5) \times 10^{-5}$ 1/year (-0.13%) | $(3 \pm 4) \times 10^{-4}$ 1/year (0.012%) |
| K | $(-860 \pm 9) \times 10^{-6}$ 1/year (-0.16%) | $(3 \pm 6) \times 10^{-5}$ 1/year (0.0059%) |
| S | $(160 \pm 6) \times 10^{-5}$ 1/year (0.018%) | $(3 \pm 5) \times 10^{-4}$ 1/year (0.0029%) |
| I | $(-310 \pm 4) \times 10^{-5}$ 1/year (-0.03%) | $(-14 \pm 3) \times 10^{-4}$ 1/year (-0.013%) |

Rate [%] was calculated as the rate fraction from the mean value.

**Figure 3:**
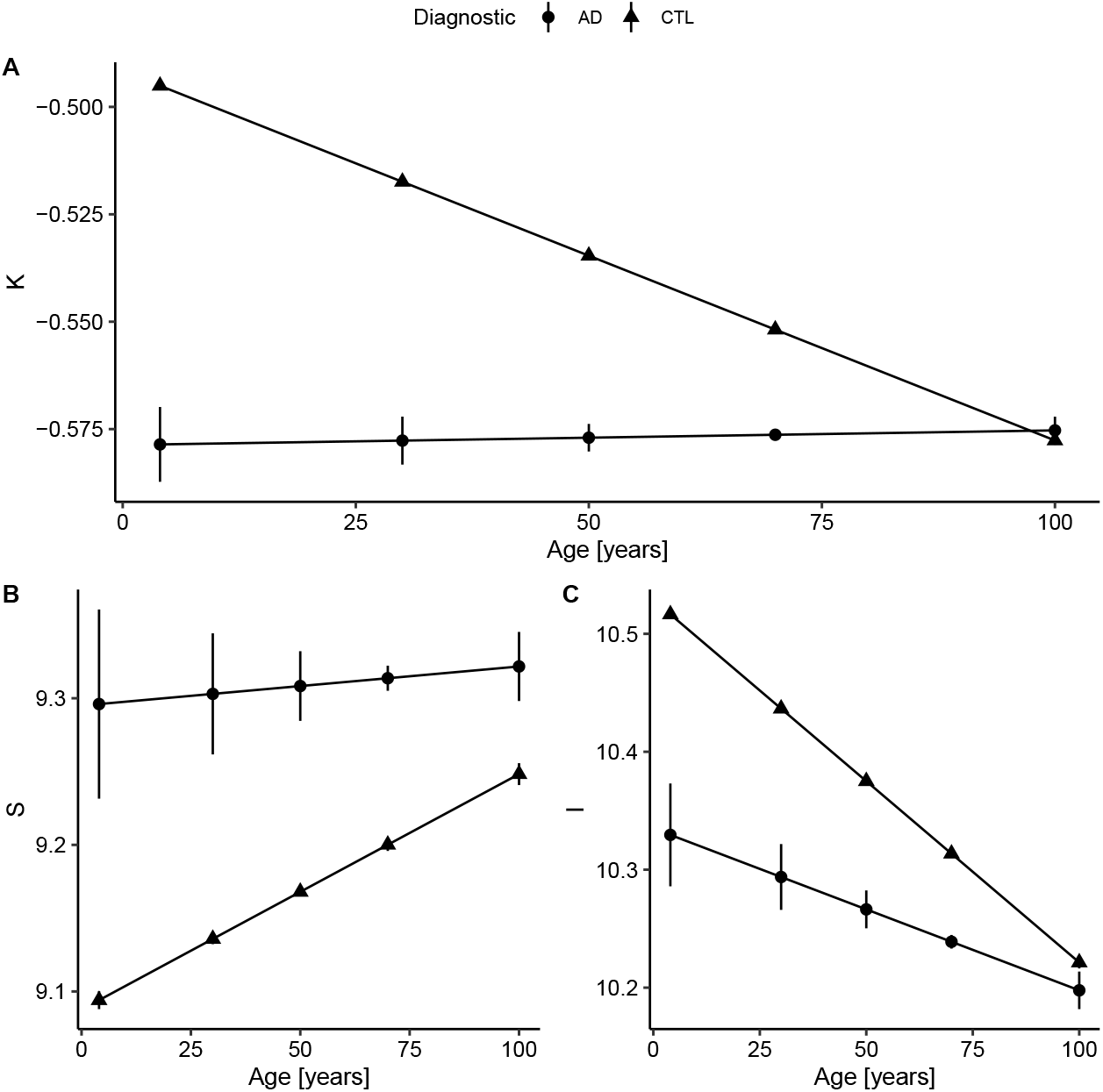
Fitted values extracted from the linear regression model after data harmonization. Bars represent the 95% confidence interval. Complete summary of linear models in Supplementary Material A, Table 5. (A) Concerning K, Alzheimer’s Disease (AD) has a shallow slope, meaning small changes with Age. Compared to healthy controls, AD values of K are almost constant and similar to older subjects (AD slope p-value < 0.0001 and CTL slope p-value < 0.0001; pairwise comparison estimate −0.00090, p < 0.0001). (B) For S, the AD and CTL patterns have similar intercepts, but statistically different slopes (AD slope p-value = 0.004 and CTL slope p-value < 0.0001; pairwise comparison estimate 0.0013, p = 0.004). (C) For I, that reflects brain volume, CTL has a decreasing volume with aging, while AD has a smaller slope (AD slope p-value < 0.0001 and CTL slope p-value < 0.0001; pairwise comparison estimate −0.0017, p < 0.0001). The curves for the AD group were extended to early ages to ease the comparison.

There is a significant difference between Control and AD slopes in K (pairwise comparison estimate −0.00090, p < 0.0001), S (pairwise comparison estimate 0.0013, p = 0.004) and I (pairwise comparison estimate −0.0017, p < 0.0001) as expected from Figure 3. For K, the AD curve is virtually flat as if it had reached a plateau, while for I, we see a constant reduction from brain isometric volume, with a reduced intercept. Comparing the mean values within AD and CTL, there is a significant difference for K (pairwise comparison estimate 0.042, p < 0.0001, representing 7.9% of the CTL mean) value, S (pairwise comparison estimate −0.14, p < 0.0001, 1.5%) and I (pairwise comparison estimate 0.11, p < 0.0001, 1.0%).

In order to complete the full description of the cortical morphology within the framework established by the Mota & Herculano-Houzel’s model, we separated the data from both Health controls and AD patients per decade and fitted the fractal dimension *α* theorized to be 1.25. The result is shown in Figure 4

**Figure 4:**
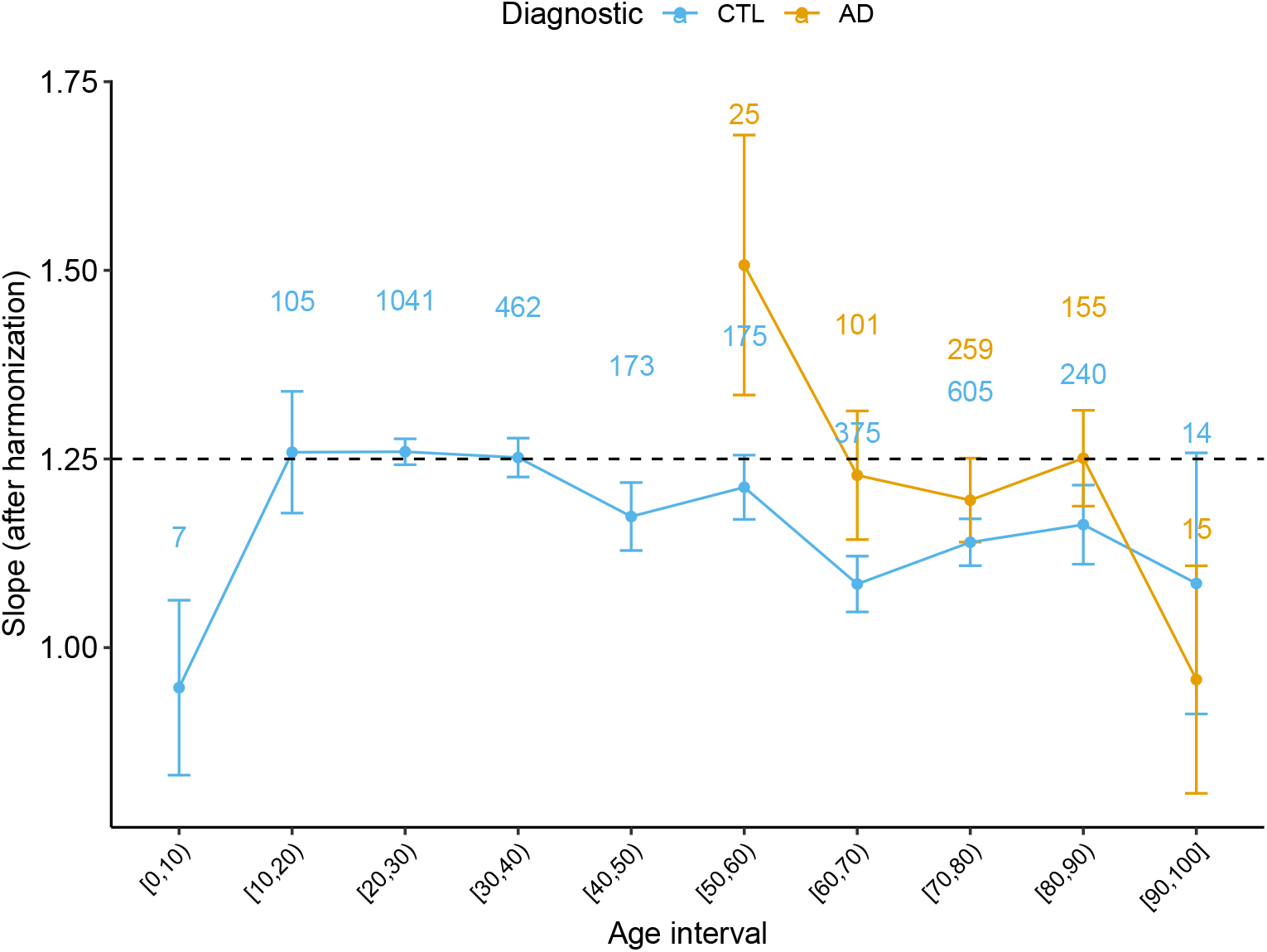
Slope *α* derived from the Cortical Folding model from Mota & Herculano-Houzel through age for Healthy Controls and AD subjects. Data points from (0, 5] and [95, 100] years old were omitted due to the reduced data with 95% Confidence Interval. The numbers on top of each point indicates the number of subjects at each interval.

As most diseases imply local or nonuniform global structural damage, we expanded the results of trajectories to the lobes. We included ROI as a fixed effect in the linear mixed model equation with harmonized variables: *Variable* ∼ *Age* × *Diagnostic* × *ROI* + (1 *Sample* : *Diagnostic* : *ROI*). Rates are summarized in Supplementary Material C. We discriminated AD and healthy aging (CTL) trends with K at the Frontal (p adj < 0.0001), Occipital (p adj < 0.0001), Parietal (p adj < 0.0001), and Temporal lobes (p adj < 0.0001), as expected from [20]. Despite the rates being smaller for AD than CTL, the results mimic the whole brain with Alzheimer’s Disease trajectories appearing as a plateau during aging. The Frontal and Parietal lobe shows the least folded pattern (K) in AD (within lobe comparison are described in Supplementary Material C). Also, there are significant differences with S CTL and AD at the Frontal (p adj = 0.00035) and Parietal lobes (p adj < 0.0001); and among I, between CTL and AD at the Frontal (p adj < 0.0001), Occipital (p adj = 0.0031), Parietal (p adj < 0.0001), and Temporal lobes (p adj < 0.0001)).

## Discussion

We analyzed a combination of datasets aiming to establish K, S, and I baseline through human lifespan. The suggested model included complexity at two levels: accounting for heterogeneous acquisition and processing and heterogeneous brain structures across healthy and disease subjects. The hypothesis is that non-homogeneous methodology (inclusion criteria, acquisition, and processing) implies more significant uncertainty due to type B errors. Thereby, we suggested a harmonization procedure that allows the comparison of data from multisite reducing this kind of experimental errors. This result is innovative by its simplicity and focuses on expanding our independent morphological variables knowledge. The main caveat of our procedure is the impossibility of accounting for all differences between the samples and using this to define a universal harmonization according to each acquisition protocol. Our method does not depend on one reference sample or a gold standard definition. The removal of the shift for each sample is carried out based on the available healthy control data, which can always be added to the analysis.

In other words, our method does not account for a possible global type B error that could shift all samples equally. By looking at the uncertainty in cortical thickness estimation, our results are compatible with previous work. Our findings of 0.02 mm (Table 2) of uncertainty in cortical thickness are in agreement with a variability less than 0.03 mm found in a test-retest analysis by Han et al. [33]. Moreover, they suggest that uncertainties of 0.15 mm across platforms of manufacturers and 0.17 mm across field strength, at this work, coupled with processing uncertainties and estimated in 0.14 mm. A most recent study from Frangou et al. [34] suggests a Mean Inter-individual variation of 0.07 ± 0.06 mm for hemisphere’s cortical thickness at all age ranges in the study compared to 0.10 mm from our findings. This compatibility suggests that the possible global type B error is close to 0; thus the harmonization could be regarded as being close to the universal one.

The typical values for the K, S, and I independent variables through the whole life span and their related uncertainties were obtained by this work. These curves not only can be used to discriminate pathological from healthy aging in cross-sectional harmonized data but also are a powerful tool to morphologically describe non-typical cortex. Estimating the uncertainties is crucial to constructing the baseline curves and was carefully developed by this work. We estimated three types of uncertainties with linear mixed models and repeated measures: type B errors (combination of multiple datasets), repeated measures, and the natural variation in the human species. The error from acquisition and processing were coupled at the individual and sample levels. We expect future works to disassociate the uncertainties in detailed factors (test-retest analysis, across platforms, field strengths, and acquisition parameters with the same sample of subjects) to better measure morphological variables of interest.

We have demonstrated the usage of the baseline curves by analyzing AD data. In terms of K, a measure of axonal tension, the AD curve is virtually flat as if it had reached a plateau of reduced brain tension. Comparing with Healthy Controls, this plateau seems to be the same experienced by the oldest subjects. Interpreting in terms of the Mota & Herculano-Houzel cortex model, AD can be regarded as a form of accelerated aging from the axonal tension perspective. On the other hand, the evolution of S, I, and even the fractal dimension *α* suggest that the AD cortex is not morphologically similar to the health one, with a very different shape, isometric volume, and time evolution. However, it is interesting to note that both S and I converge to a value compatible with the oldest health subjects. In the future, correlating these results with the elasto-mechanic properties of the AD cortex will help to understand better the physical limits of Mota & Herculano-Houzel model and expand it theoretically.

A significant result that could only be achieved by combining multiple data is the value of the fractal dimension *α* through human lifespan and its evolution through atypical conditions. We extended the analysis in de Moraes et al. [35] and verified how the slope *α* behaves with healthy aging. We found that the slope is compatible with the theoretical prediction for subjects between 20 and 60 years old. Defining this range of applicability of the theory is essential to understand the period of life in which the model’s basic assumptions are still valid. After, we confirmed the findings that the slope escapes the model after 60 years old, diverging from the theoretical value of 1.25. Before 15 years old, we have found a hint that the slope is also inferior to 1.25, in disagreement with the model. Along, a non-linear pattern is suggested by the K baseline function before 20 years. One of the leading hypotheses to explain the deviation of the measured *α* from the expected value in both cases (before 15 and after 60 years old) is the breakdown of the homogeneity on the cortical surface. The suggestion of a homogeneity breakdown makes it particularly interesting to further study AD in the age range compatible with 1.25. Understanding the regimes outside the model is a fundamental step towards the theoretical understanding of cortical folding. Coupled with analysis of neuropathologies such as AD, these empirical results improve the cortical folding theory.

More than discriminating disease and healthy subjects, understanding the pathology trajectory is vital to defining landmarks, indicating future injury, and collaborating to develop efficient treatment. Notably, in Alzheimer’s Disease and its prodromal form, Mild Cognitive Impairment, subsequent phases of damage may start at least 20 years before the diagnostic [36]. The structural damage is an initial state than cognitive alterations, as memory and clinical functions [37]. As pointed out by Fjell et al. [38], healthy aging also contributes to reductions in cortical thickness, and understanding the overlap between healthy aging and pathological aging triggers is crucial to segregate both events. Nevertheless, the possible similarity of structural damage in Alzheimer’s Disease and aging is explored by multiple previous studies [13, 39–41]. We hypothesize that the structural changes due to AD are reflected in measures of the brain’s global structure and shape. Therefore, cortical folding derived variables K, S, and I are sensitive to this perturbation. We verified this hypothesis by analyzing the difference in aging trajectories between healthy and pathological brains.

At a more refined grain scale, the lobes, the results are compelling to confirm [13] suggestion that K, S, and I together would be a powerful tool to discriminate similarly but distinct events, as AD and aging. In this case, the difference between events is in the cortical structure’s changing velocity or rate. Then with the rates for K, S, and I at the lobes, we could explore whether a lobe would unfold faster. For AD, the temporal lobe has the biggest unfolding (K) and shrinking rate (I), while healthy aging is more aggressive at the Parietal lobe, followed by the temporal and frontal lobes.

The natural expansion of this work is to include Mild Cognitive Impairment (MCI) subjects. As seen before, the MCI is an intermediary step between healthy aging and AD in the disease progression, cognitive impairment, and structural changes [20, 36, 37]. Also, we intend to continue this work by applying this approach to a longitudinal dataset composed of MRI structural images with MCI subjects that eventually convert to AD and compare to the cortical folding components of non-converters. Including this data would allow having a more reasonable aging trajectory by having the transition period from healthy aging to pathological aging. Further, we expect to explore K, S, and I function with age for the age range comprising newborns to 25 years old in future studies. This study will help better understand the value of the slope *α* at this age, revealing if there is an inflection point that can be connected to the development/aging transition in human brains [42], such as suggested by the data.

This work successfully achieved its goal of combining multiple samples with a simplistic harmonization procedure that adjusts K, S, and I values for the 3650 subjects. Hereafter we estimated K, S, and I typical values and aging rates to discriminate against Alzheimer’s Disease subjects based on their changing rates. To validate the proposed methodology, we analyzed results for Cortical Thickness, which are in concordance with previously published findings. At a glance, the significant differences found in aging rates for the cross-sectional data are suggestive that a brain with Alzheimer’s Disease has premature and accelerated aging in terms of morphology.

## Data Availability

Data from previous publications are available from their repositories: Wang et al., 2016 and Wang et al., 2019. Data from AHEAD and AOMIC datasets with the cortical folding variables extracted by these authors are available.
Data acquired at IDOR does not have clearance for public sharing patients' information that could lead to identification due to local Ethics Committee approval restrictions but will be shared upon request.

https://zenodo.org/record/5750619

https://www.pnas.org/doi/pdf/10.1073/pnas.1610175113

https://zenodo.org/record/2626779

## Acknowledgements

This work was funded by the Research Support Foundation of the State of Rio de Janeiro (FAPERJ), National Council for Scientific and Technological Development (CNPq), and intramural grants from Instituto D’Or de Pesquisa e Ensino (IDOR). Bruno Mota is supported by Instituto Serrapilheira (grant Serra-1709-16981) and CNPq (PQ 2017 312837/2017-8). We are thankful to the research team and volunteers for participating in this research project. The authors have no competing interests.

## Author Contributions

F.H.P.M., V.B.B.M, and B.M. designed research; F.H.P.M., V.B.B.M, F.T.M., and B.M. performed research; F.H.P.M., V.B.B.M and B.M. analyzed data; and F.H.P.M., V.B.B.M and B.M. wrote the paper. All authors reviewed the manuscript.

## Supplementary Material

**Table 4:** Summary for each dataset. Mean value ± standard deviation. Cross-sectional analyses were performed compilating all samples into one heterogeneous dataset.

| Datasets | Diagnostic | N (F) | Age [years] | MRI equipment | FreeSurfer |
| --- | --- | --- | --- | --- | --- |
| ADNI [23] | CTL | 868 (445) | 75 ± 6.5 | multiple 3T | v5.3 |
| | AD | 542 (241) | 75 ± 8 | | |
| AHEAD<sup>1</sup> [21] | CTL | 100 (56) | 42 ± 19 | Philips Achieva 7T | v6 |
| AOMIC PIOP01 [22] | CTL | 208 (120) | 22 ± 1.8 | Philips Achieva 3T | |
| AOMIC PIOP02 [22] | CTL | 224 (128) | 22 ± 1.8 | Philips Achieva dStream 3T | |
| HCP900r<sup>1</sup> [24] | CTL | 881 (494) | 29 ± 3.6 | Siemens Skyra (modified) 3T | v5.3<sup>2</sup> |
| IDOR [20] | CTL | 77 (53) | 66 ± 8.4 | Philips Achieva 3T | v6 |
| | AD | 13 (8) | 77 ± 6.1 | | |
| IXI-Guy's<sup>3,4</sup> | CTL | 314 (175) | 51 ± 16 | Philips Intera 3T | v5.3 |
| IXI-HH<sup>4</sup> | CTL | 181 (94) | 47 ± 17 | Phillips Gyroscan Intera 1.5T | v5.3 |
| IXI-IOP<sup>6</sup> | CTL | 68 (44) | 42 ± 17 | GE 1.5T | v5.3 |
| NKI [25] | CTL | 168 (68) | 34 ± 19 | Siemens Magnetom 3T | v5 |
| OASIS<sup>7</sup> [26] | CTL | 312 (196) | 45 ± 24 | Siemens Vision Scanner 1.5T | Dev 20061005 |

Diagnostic: CTL – Healthy control; AD – Alzheimer's Disease
<sup>1</sup>AHEAD and HCP900 datasets provided an age range for each subject. Therefore, we supposed that each subject's age was the provided interval's mean age.
<sup>2</sup>modified version
<sup>3</sup>http://brain-development.org/ixi-dataset/
<sup>4</sup>IXI subset: Guy's Hospital
<sup>5</sup>IXI subset: Hammersmith Hospital
<sup>6</sup>IXI subset: Institute of Psychiatry
<sup>7</sup>Data were only available for hemispheres analysis, not used for the lobes tests.

**Table 5:**
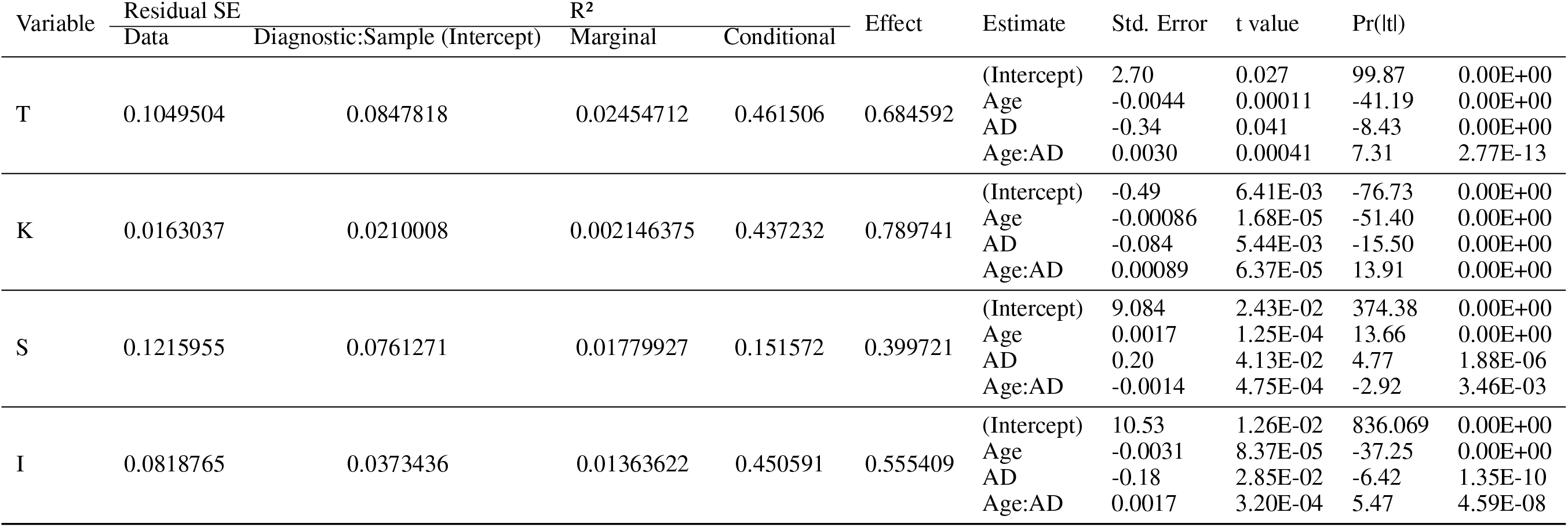
Linear mixed models summary. Raw data, with no harmonization. 4364 observations; 11 Diagnostic:Sample; 8 Sample. Equation: *Variable* ∼ *Age × Diagnostic* + (1|*Sample*: *Diagnostic*).

### A Supplementary Tables, Figures and Results

### B Bayesian model selection

In order to show that the harmonization procedure used were the most suitable, one can compare it with other models to describe the data. For example, the angular coefficient could also be a random effect for the LMM, varying across the samples. Using Bayesian model selection [43], one can show that the LMM used in this work is the most probable description to the data.

Considering our dataset comprised by *J* samples where 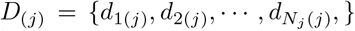 denotes the data of a morphological variable in the *j*-th sample. The degree of belief in a model *M* that describes a normally distributed data for each age is given by the posterior probability:

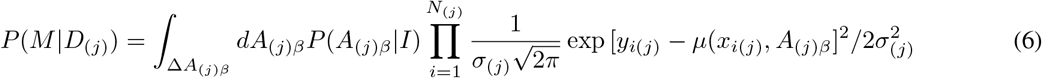

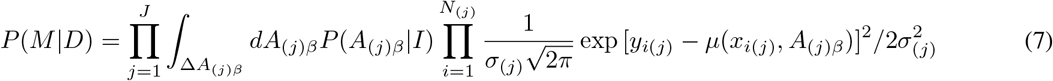

The term *µ*(*x*_*i*(*j*)_, *A*_(*j*)*β*_) is given by the model *M* proposed to describe the data, with the set {*A*_(*j*)*β*_} being the free parameters of the model defined within a range Δ*A*_(*j*)*β*_. A prior probability *P* (*A*_(*j*)*β*_|*I*) is assigned for all free parameters reflecting the prior knowledge about their values. The basic model of this work assumes that the log of morphological variables follow *µ* = *ax* + *b*_(*j*)_, with the dispersion *σ*_(*j*)_ for each samples. Uninformative priors were used for the free parameters. This integral can be easily done numerically using Python3 with PyMC3 package or similar. The advantage of this approach is that it naturally penalizes the insertion of unnecessary new parameters avoiding over-fitting.

With multiple models *M*_*i*_, one defines the odds as 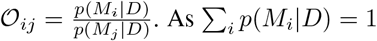, the probability of the model *M*_1_ is given by:

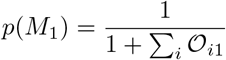

The alternative models considered by this work were: i) *µ* = *ax*+*b*_(*j*)_, with the same dispersion *σ*_(*j*)_; ii) *µ* = *a*_(*j*)_*x*+*b*_(*j*)_, with the dispersion *σ*_(*j*)_ for each sample; iii) *µ* = *a*_(*j*)_*x* + *b*_(*j*)_, with the same dispersion *σ*_(*j*)_ for each sample; iv) We also tested the simplest non-linear trend *µ* = *cx*^2^ + *ax* + *b*_(*j*)_, with the dispersion *σ*_(*j*)_ for each sample; We have found that the LMM used in the work is the most probable. The code implementing this approach is given by the authors.

It is important to note the all parameters calculated using the LMM approach can be obtained in this framework by a slight modification of equation 7. Instead of marginalizing over all the parameters, by left one out, we obtain *P* (*A*_*j*_1|*D*) which gives directly the most probable value for the parameter *A*_*j*_1 and its credible region.

### C Expanded results to lobes

We expanded the analyses of trajectories to the lobes by including ROI as a fixed effect in the linear mixed model equation: *Variable* ∼ *Age* × *Diagnostic* × *ROI* + (1|*Sample* : *Diagnostic* : *ROI*). Rates are summarized in Table 6 and Figure 5. Differences within lobes and rate differences within diagnostics are described in Figure 6.

**Table 6:**
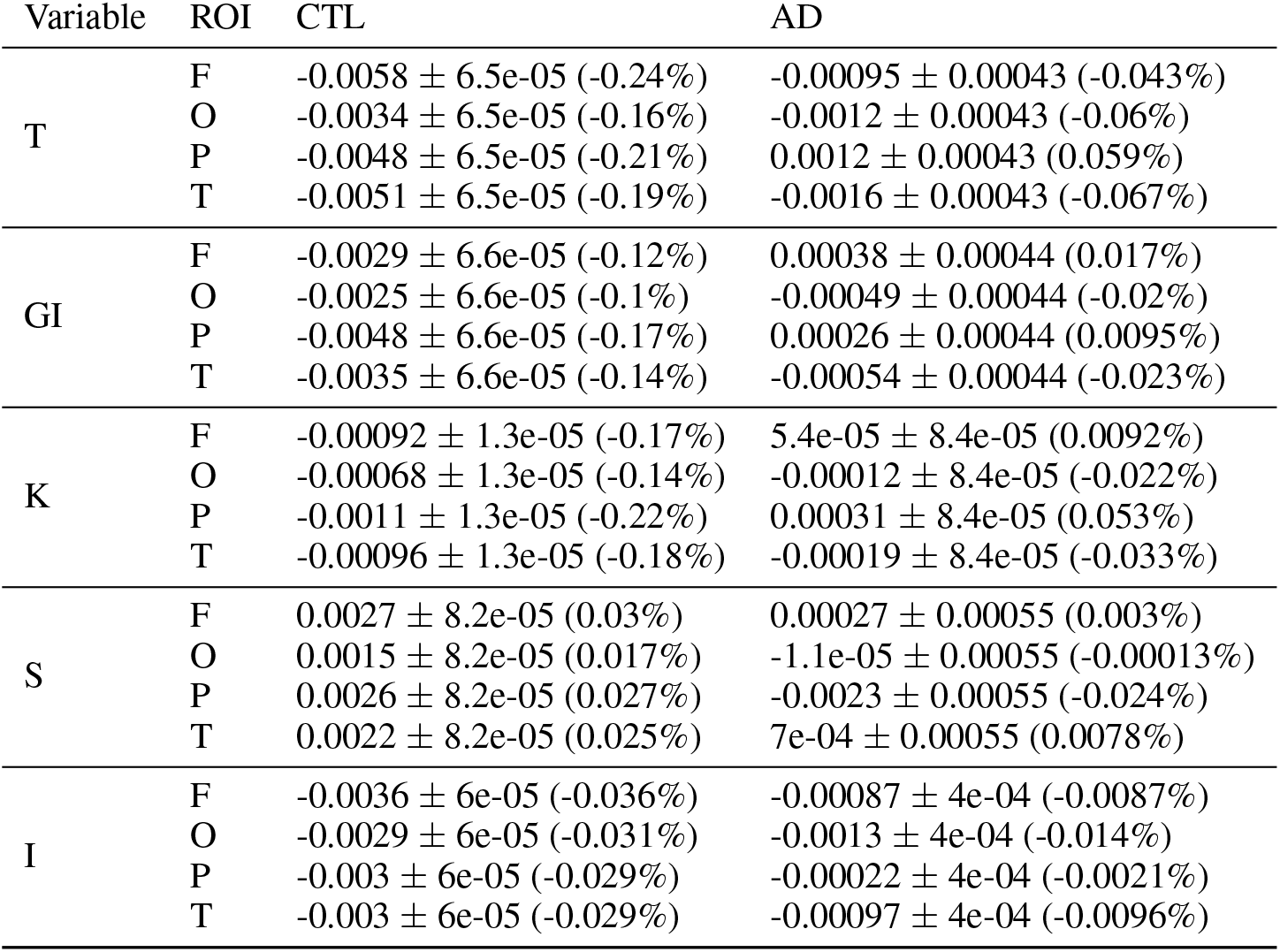
Changing rate per year for each variable, diagnostic and lobe. Mean value *±* standard deviation.

**Figure 5:**
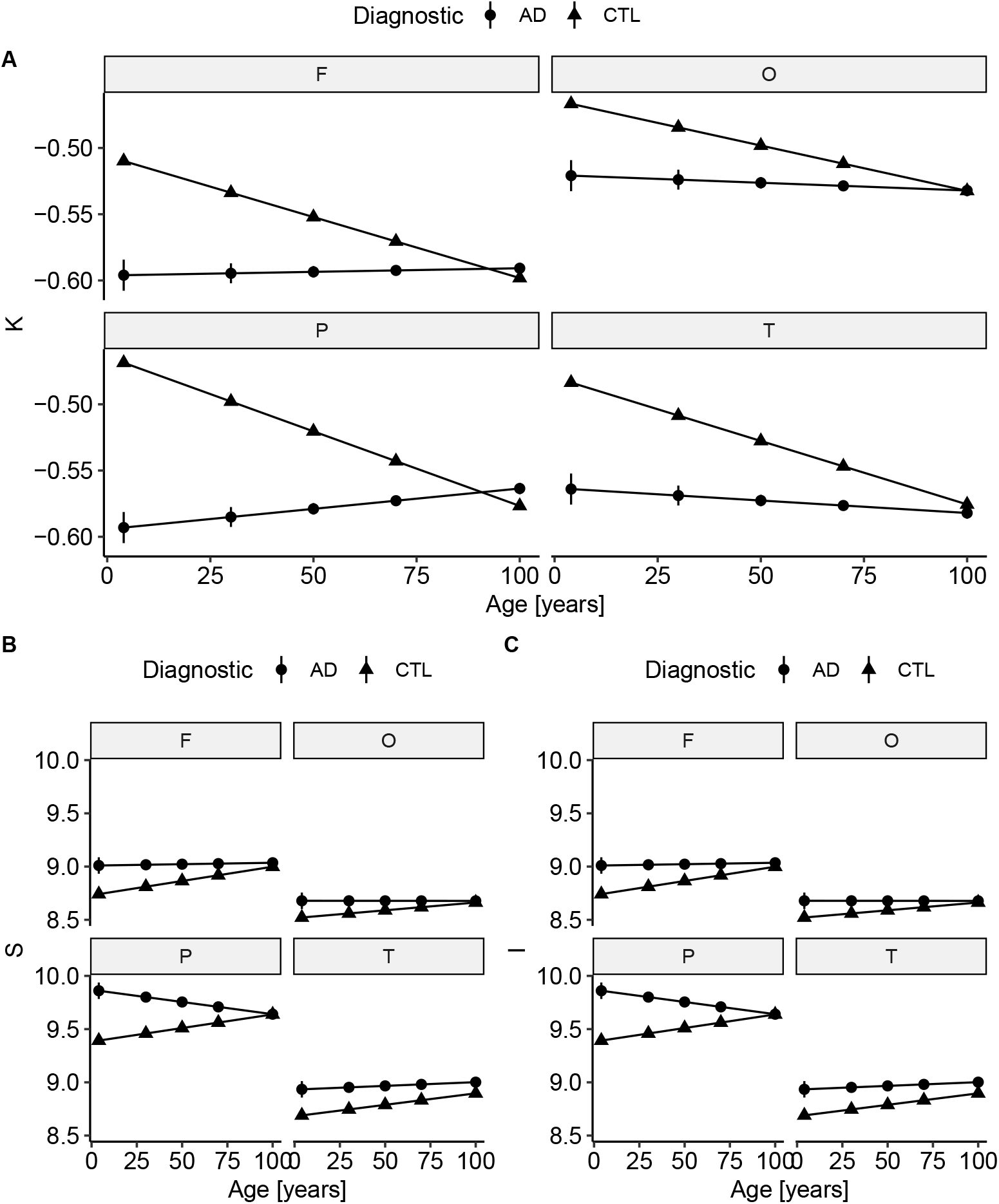
Fitted values for (A) K, (B) S and (I) and age for all for the Frontal lobe (“F”), Parietal lobe (“P”), Occipital lobe (“O”) and Temporal lobe (“T”) after data harmonization. Bars represents 95% confidence interval.

**Figure 6:**
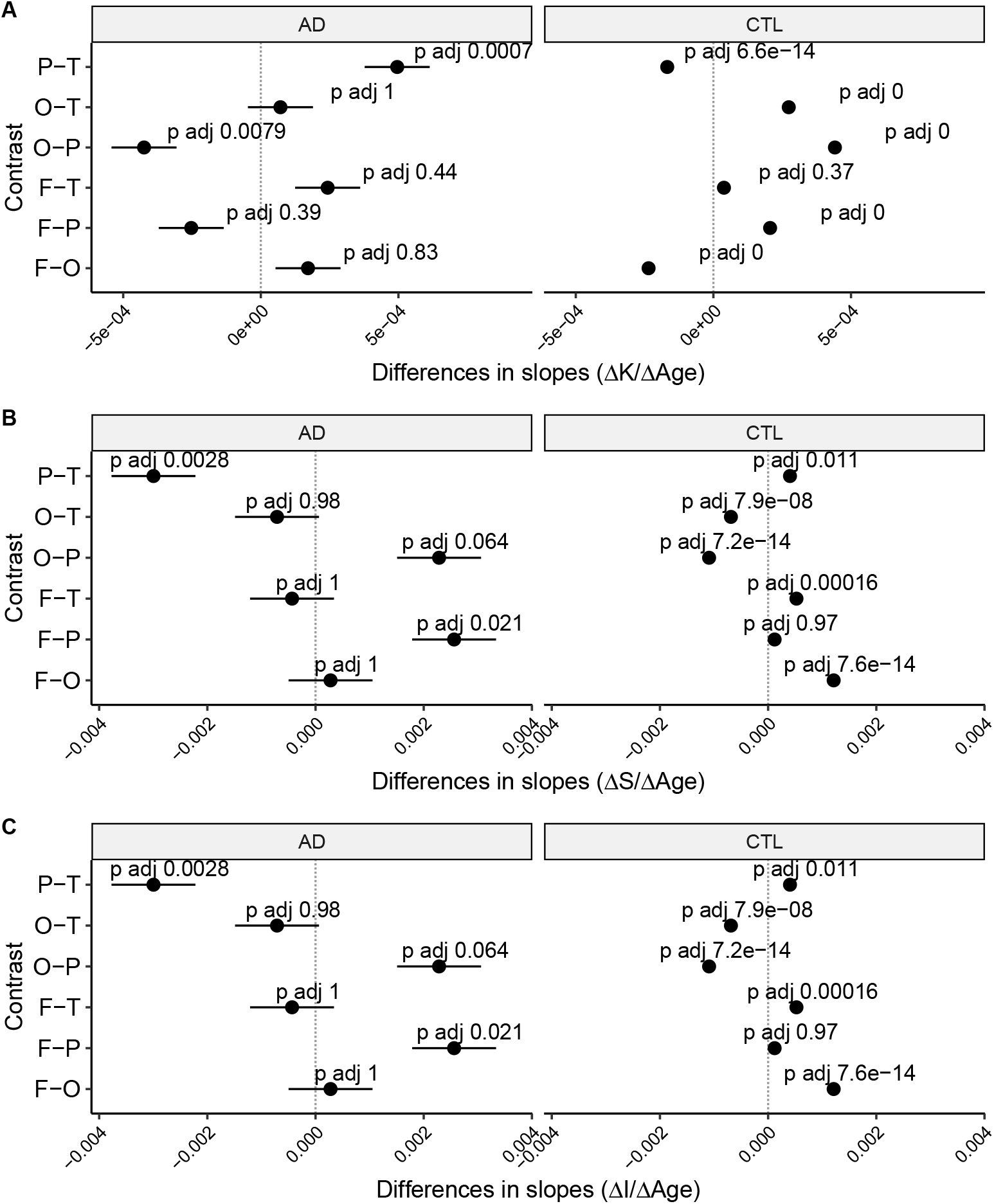
Difference in slopes for each lobe within diagnostics after data harmonization. Bars represent a 95% confidence interval. (A) K, (B) S, and (C) I.

#### C.1 Repeated measurements

The random error is an essential fraction of the global uncertainties, as it is the variance that occurred after multiples measures in the same condition. To estimate the reproducibility within subjects, we processed (as described in Materials and Methods) the first 50 subjects with all three T1w images from the AOMIC ID1000 [22] dataset (described in Supplementary Material Table 7). The AOMIC ID1000 comprises multiple Magnetic Resonance Imaging protocols, especially three structural T1w images acquired during the same scan with the same protocol in a Philips Intera 3T. The select sample has the advantage of being composed of only healthy human subjects within a narrow age range, from 20 to 26 years old. We hypothesized that should be no difference of means within the three runs, despite the variation for each subject (Figure 7).

**Table 7:** Summary for the 50 subjects of AOMIC ID1000 dataset. Mean value *±* standard deviation.

| Session | Age [years] | T | K | S | I |
| --- | --- | --- | --- | --- | --- |
| 1 | $23 \pm 1.7$ | $2.7 \pm 0.093$ | $-0.51 \pm 0.017$ | $9.1 \pm 0.093$ | $10 \pm 0.078$ |
| 2 | $23 \pm 1.7$ | $2.7 \pm 0.093$ | $-0.51 \pm 0.017$ | $9.1 \pm 0.095$ | $10 \pm 0.076$ |
| 3 | $23 \pm 1.7$ | $2.7 \pm 0.094$ | $-0.51 \pm 0.017$ | $9.1 \pm 0.091$ | $10 \pm 0.078$ |

**Figure 7:**
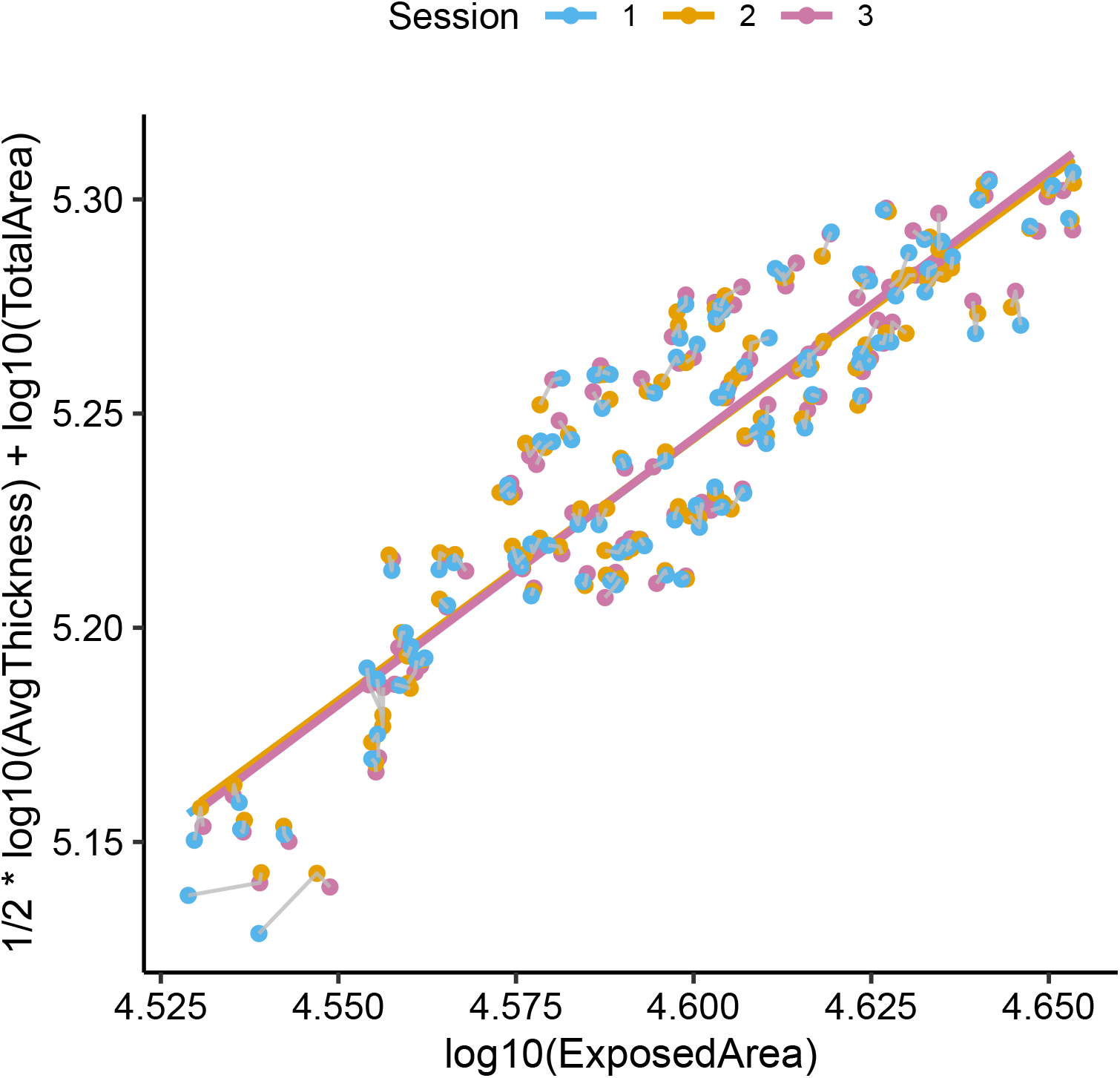
Cortical folding model with the traced path for each hemisphere of each subject (gray line). The sample respects the model with slope *α* = 1.23 *±* 0.06, 95% confidence interval = (1.12,1.34).

To estimate the uncertainty/variation of repeated measures, one could estimate the distribution of standard deviations of the data. We calculated the standard deviation for each subject and hemisphere across the three images, for the Cortical Thickness, K, S, and I.

We compared the means for each run with a Repeat Measure ANOVA. There is no significant difference of means through runs (1, 2 and 3) (Figure 8): Cortical Thickness [mm], F(2, 198) = 0.481, p = 0.62, eta2[g] = 0.000192; GI, F(2, 186) = 0.196, p = 0.81, eta2[g] = 0.00001; K, F(2, 198) = 0.138, p = 0.87, eta2[g] = 0.0000144; S, F(2, 185) = 0.352, p = 0.69, eta2[g] = 0.0000798; I, F(2, 198) = 0.775, p = 0.46, eta2[g] = 0.0000523. The estimate standard deviations are: Cortical Thickness [mm], 0.019 *±* 0.013 mm; GI, 0.0069 *±* 0.0047; K, 0.0017 *±* 0.0012; S, 0.014 *±* 0.01; I, 0.0064 *±* 0.0043 (Figure 9).

**Figure 8:**
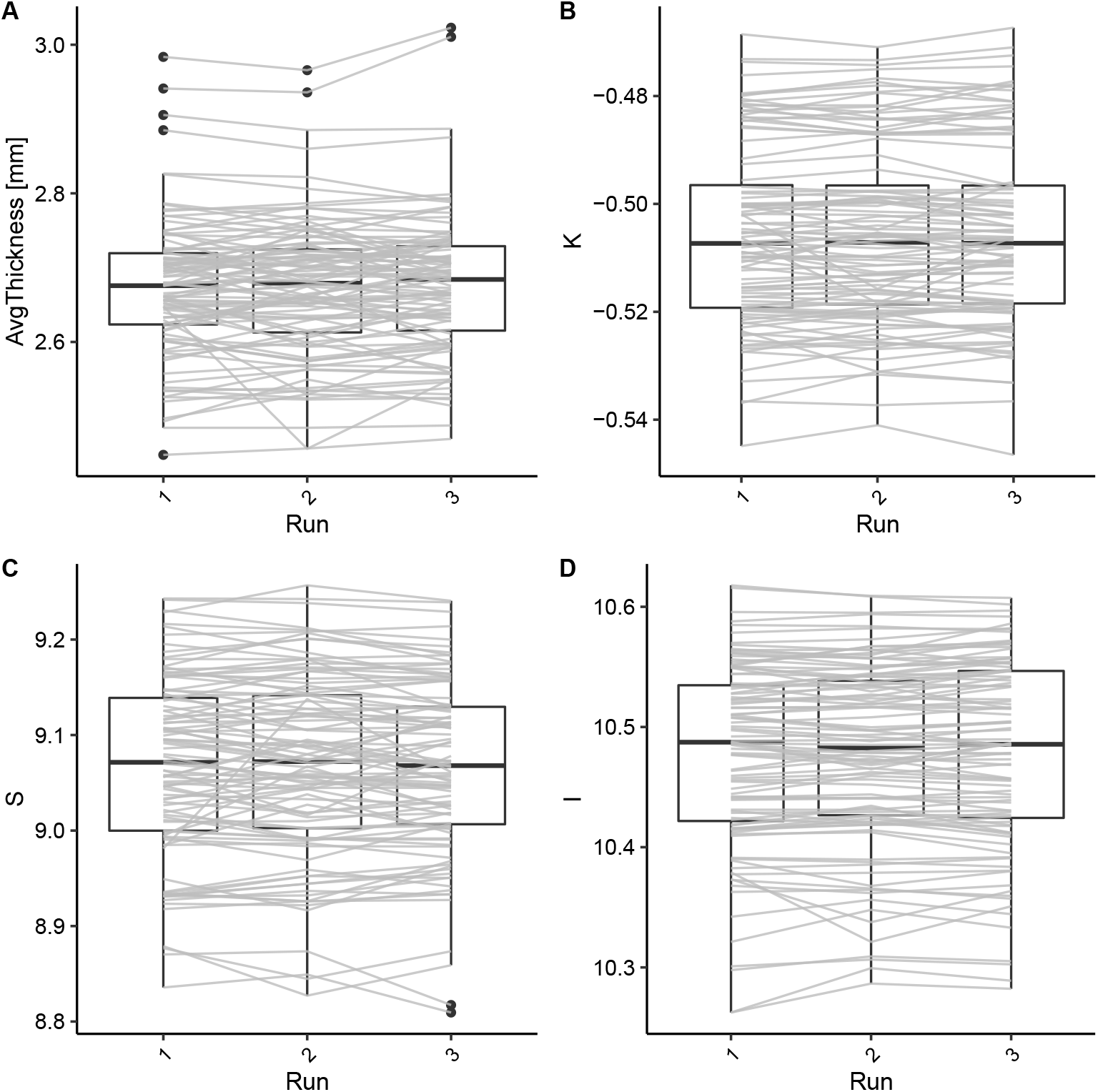
Trajectories for each hemisphere of each subject through the acquisition runs. There is no significant difference in means. (A) Cortical Thickness [mm]: F(2, 198) = 0.481, p = 0.62, eta2[g] = 0.000192; (B) K: F(2, 198) = 0.138, p = 0.87, eta2[g] = 0.0000144; (C) S: F(2, 185) = 0.352, p = 0.69, eta2[g] = 0.0000798; (D) I: F(2, 198) = 0.775, p = 0.46, eta2[g] = 0.0000523.

**Figure 9:**
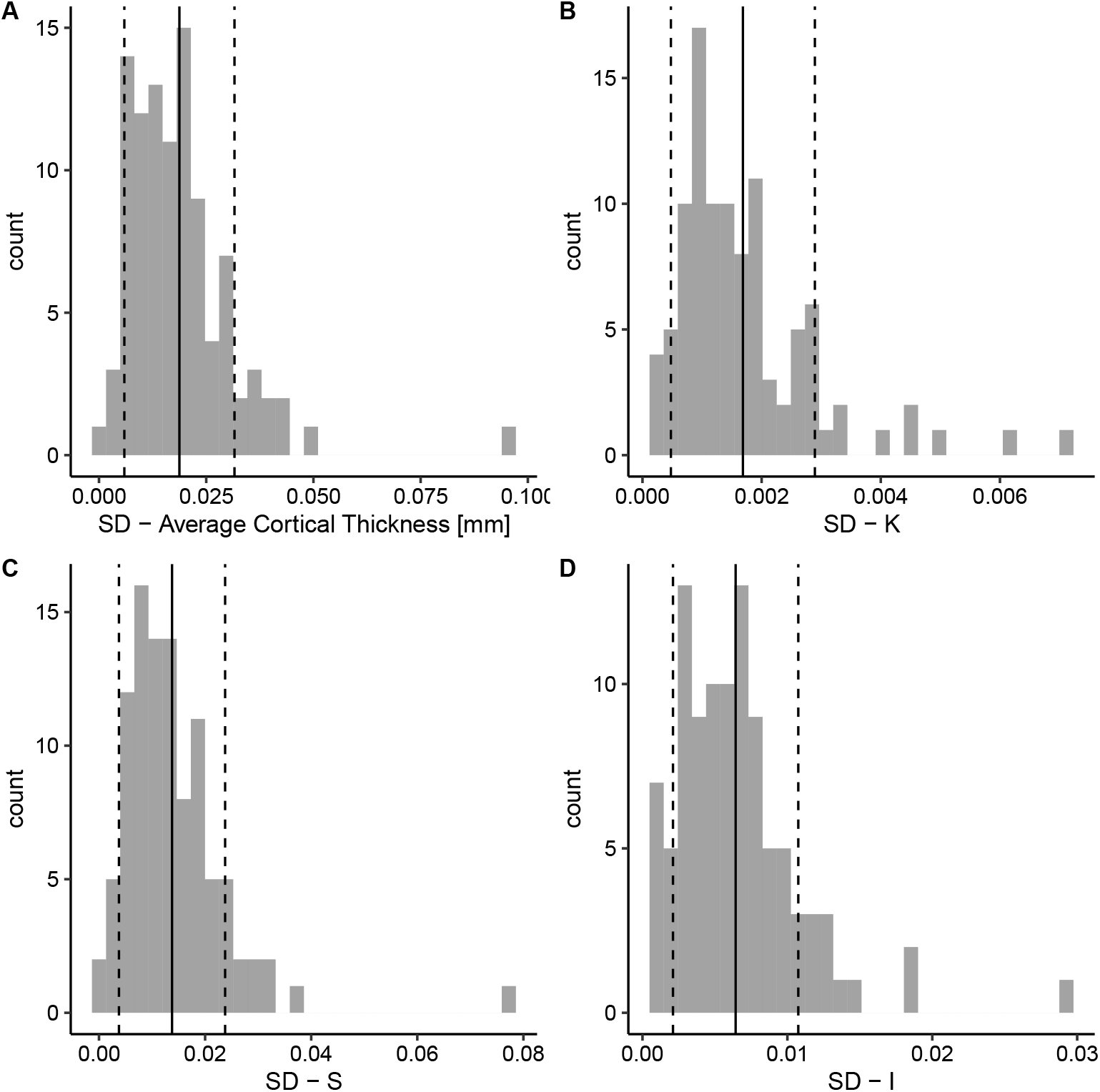
Standard Deviation (SD) distribution for each variable. The solid line represents the mean value, and the dashed line the standard deviation of the distribution. (A) Cortical Thickness: 0.019*±*0.013 mm; (B) K: 0.0017*±*0.0012; (C) S: 0.014*±*0.01; (D) I: 0.0064*±*0.0043.

These results suggest high reliability in subsequently acquired images processed with FreeSurfer v6.0, in concordance with [44] finds for reliability in FreeSurfer processing comparing volumes measurements. We suggest that the estimated standard errors accurately estimate the uncertainty due to a repeated acquisition and image processing within the same site and parameters.

### D multicenter influence on cortical folding model and its independent morphological components

We estimated the influence of multicenter datasets in estimating K, S, and I typical values and rates per year across the human lifespan, including as random effects at the linear mixed model. Using these models allows one to calculate the residual variance arising from the methodological heterogeneity and the variance from each category, the different included data samples in this manuscript.

The systematic shifts, estimated from the Linear Mixed Models (LMM, *Variable* ∼ *Age* × *Diagnostic* × *ROI* + (1|*Sample* : *Diagnostic* : *ROI*)), can be used as inputs to multisite harmonization procedures, reducing the influence of heterogeneous methodology (Figure 1). The harmonization consists of subtracting the estimated shift from the raw data of each sample (*X*_*harmonized*_ = *X*_*raw*_ − *shift*_*X*_). It reduces the systematic error due to the samples, and consecutively, the total uncertainty of the variable (Table 8). A step-by-step code is available [35].

**Table 8:** Comparison of estimated error after the multisite harmonization.

| Variable | Natural Fluctuation | $\sigma_{acquisition} + \sigma_{processing}$ | | $\sigma_X = \sqrt{\sum \sigma_i^2}$ |
| --- | --- | --- | --- | --- |
| | ($\sigma_{withinspecienaturalvariation}$) | random<sup>1</sup> | systematic ($\sigma_{sample}$) | |
| T [mm] | 0.1 | 0.019 | 0.000 | 0.10 (4.10%) |
| $A_T$ [mm<sup>2</sup>] | 9700 | 290 | 0 | 9704.33 (9.63%) |
| $A_E$ [mm<sup>2</sup>] | 2900 | 70 | 0 | 2900.84 (7.5%) |
| GM volume [mm<sup>3</sup>] | 33000 | 3600 | 0 | 33195.8 (13.27%) |
| GI | 0.097 | 0.0069 | 0.000 | 0.97 (3.74%) |
| K | 0.017 | 0.0017 | 0.000 | 0.017 (3.19xx%) |
| S | 0.12 | 0.014 | 0.000 | 0.12 (1.32%) |
| I | 0.082 | 0.0064 | 0.000 | 0.082 (0.79%) |
| <sup>1</sup>repeat measures, mean standard deviation |  |  |  |  |

Future studies can progress on the limitation by detailing influences and increasing specificity in methodology, such as the scan field strength, model and manufacturer, and decoupling acquisition imaging processing components.

## Footnotes

1 One may understand this as similar to “Principal Component Analysis”, a change of basis on the data representation, but using scaling considerations rather than covariance eigenvectors to define the principal directions. The new set of variables are combinations of the usual geometrical variables used to describe the cortex. The principal component K is given by the power-law relation, which contains the correlation between the variables. The variables S and I are orthonormal to K.

2 Structural T1w images from AOMIC ID1000 dataset, described in Material and Methods.

